# Comparing measured and calculated blood loss after a caesarean birth

**DOI:** 10.64898/2026.07.16.26358295

**Authors:** Raoul Mansukhani, Monica Arribas, Nike Bello, Rizwana Chaudhri, Amber Geer, Katharine Ker, Projestine Muganyizi, Daniele Prowse, Ian Roberts

**Affiliations:** Global Health Trials Group, London School of Hygiene & Tropical Medicine, London, UK; Department of Obstetrics and Gynaecology, University of Ibadan College of Medicine, Ibadan, Nigeria; Global Institute of Human Development, Shifa Tameer-e-Millat University, Islamabad, Pakistan; University of Dar es Salaam Mbeya College of Health and Allied Sciences, Dar es Salaam, Tanzania

**Author notes:** Corresponding author: Raoul Mansukhani, Global Health Trials Group, London School of Hygiene & Tropical Medicine, London, WC1E 7HT.

## Abstract

**Introduction:** Accurate measurement of blood loss after caesarean birth is important for early identification of postpartum haemorrhage. Commonly used visual estimation underestimates blood loss. We compared two methods for objectively assessing blood loss.

**Methods:** Measured blood loss was obtained by weighing swabs and pads combined with blood from suction and drapes. Calculated blood loss was derived from maternal weight and peripartum haemoglobin change. Data were from the I’M WOMAN trial investigating tranexamic acid for postpartum haemorrhage prevention. This was a secondary analysis of prospectively collected trial data. We reported median (IQR) blood loss, assessed agreement using Bland-Altman analysis and compared the percentages of women exceeding 500 ml, 1000 ml and 1500 ml. We graphed proportional haemoglobin drop by categories of measured blood loss, stratified by anaemia status. We compared AUCs for measured and calculated blood loss predicting haemodynamic compromise (shock index above 1.0) and death or near-miss.

**Results:** A total of 10,393 women were included in this study. Median measured and calculated blood loss were 545 ml (IQR 430-700) and 505 ml (IQR 180-908) respectively, with weak correlation (Spearman’s rho=0.28). The IQR was wider for calculated blood loss than for measured blood loss, indicating greater variability. Bland-Altman analysis showed a small mean bias of -39 ml but wide limits of agreement (lower -1196 ml, upper 1118 ml). Measured versus calculated blood loss exceeded 500 ml in 60% versus 51% of women, 1000 ml in 7% versus 21%, and 1500 ml in 2% versus 8%. Women without anaemia had a greater proportional haemoglobin drop than women with anaemia for blood losses below 1000 ml. Measured blood loss had better predictive ability for death or near-miss (AUC 0.87 vs 0.76, p<0.001) and haemodynamic compromise (AUC 0.66 vs 0.62, p=0.001).

**Conclusion:** While median measured and calculated blood losses were similar, they were only weakly correlated. Calculated blood loss classified more women as having blood loss above higher thresholds. Women without anaemia had a greater proportional haemoglobin drop than women with anaemia for blood loss below 1000 ml. Measured blood loss had a modest advantage in predicting death or near-miss and haemodynamic compromise, but calculated blood loss remains an objective measure suitable as a trial outcome when direct measurement is not feasible.

## Introduction

Haemorrhage is the leading cause of maternal death.^1^ More than 85% of all maternal deaths occur in sub-Saharan Africa and South Asia.^2^ Most maternal deaths due to haemorrhage are preventable.^3,4^ Clinicians typically estimate maternal blood loss visually despite visual estimation being known to underestimate actual blood loss.^3,5,6^ Underestimation of blood loss may result in a woman receiving treatment late or not at all.^6^ Prompt diagnosis and treatment of postpartum haemorrhage (PPH) are crucial for preventing death and near-miss.^6^ The WHO has recently updated its guidelines highlighting the importance of early diagnosis and now recommends that objective techniques be used for measuring blood loss.^7,8^

Measured blood loss has been proposed as an objective method for measuring blood loss during birth.^4,9^ For caesarean births, it is obtained by quantifying blood from suction canisters, swabs, gauzes and pads during surgery and from calibrated drapes placed beneath the woman following surgery.^4^ By contrast, calculated blood loss is often used as an outcome measure in studies of obstetric bleeding.^10–14^ Calculated blood loss is obtained using three measurements: maternal weight and prebirth and 24-hour postpartum haemoglobin.^15^

A recent study compared measured and calculated blood loss in women having a vaginal birth in France.^9^ We compared measured and calculated blood loss in women having a caesarean birth in Nigeria, Pakistan and Tanzania.

## Methods

### Study participants

We used data from the I’M WOMAN trial, an ongoing double-blind randomised controlled trial comparing the efficacy and safety of intramuscular (IM) tranexamic acid, intravenous (IV) tranexamic acid and placebo for the prevention of PPH and other adverse maternal outcomes in women at increased risk of PPH.^16,17^ Women aged 18 years or older admitted to hospital for a vaginal or caesarean birth with one or more risk factors for postpartum haemorrhage were eligible for inclusion (Appendix 1). Women with a known allergy to tranexamic acid were excluded. Eligible women were randomised to receive: a) 1 gram of tranexamic acid as two 5 ml IM injections (100 mg/ml) and IV placebo (10 ml 0.9% sodium chloride); b) 1 gram of tranexamic acid by IV injection and two 5 ml IM placebo injections; or c) matching placebo. Sites were selected based on a network established through previous obstetric trials.^18,19^

I’M WOMAN trial recruitment started on 22 April 2024. A protocol amendment was made on 30 August 2024 instructing sites to measure women’s prebirth and postpartum venous haemoglobin and weight. This study includes women recruited from sites where this amendment had been implemented.

The I’M WOMAN trial protocol was approved by the London School of Hygiene & Tropical Medicine’s Ethics Committee (reference 28252). A member of the site trial team identified potentially eligible women and approached them with the agreement of the primary carer. Women were given information about the trial in a language they understood. The site trial team explained the purpose of the trial, that participation would not involve any change to the woman’s birth plan, and that she would receive all usual interventions for preventing PPH and any other care needed. Women were informed that participation was voluntary and that choosing not to take part would not affect their care. Women who agreed to take part provided written consent. If a woman was unable to read or write, the participant information sheet was read to her and she marked the consent form with a cross or thumbprint. In this situation, an impartial witness signed the consent form to confirm the woman’s mark. The consent procedures are described in detail in the study protocol.^16^

### Procedures and outcomes

Information about the woman’s health and pregnancy status was assessed as part of trial entry. Women were weighed after admission and before giving birth. If scales were unavailable or if measurement was not possible then weight was estimated by a clinician. We measured haemoglobin from venous blood samples taken after admission but before birth, and at 24 hours after birth or discharge, whichever was sooner. Blood pressure and heart rate were measured at regular intervals after birth according to each site’s usual standard of care, with the lowest blood pressure and corresponding pulse rate recorded up to 24 hours after birth or at discharge, whichever occurred first.

Measured blood loss was obtained from: 1) blood collected by suction during surgery, with amniotic fluid estimated and deducted from the suctioned volume; 2) blood absorbed by swabs, pads, gauzes and cloths during surgery, weighed before and after use (1g = 1ml blood); and 3) blood collected in calibrated drapes placed beneath the woman immediately after surgery for 1 hour (extended to 2 hours only if heavy bleeding continued), followed by visual estimation for up to 24 hours after birth.

Calculated blood loss was defined as described by Shook and colleagues as the product of a woman’s estimated blood volume and her proportional peripartum change in haemoglobin (equation 1).^15^ Blood volume (litres) was estimated by multiplying the woman’s weight (kg) by 0.085 (equation 2).^15^ In our calculations, we reduced postpartum haemoglobin by 10 g/L for each unit of red blood cells transfused.^20,21^

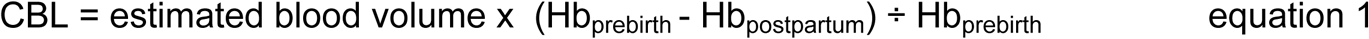

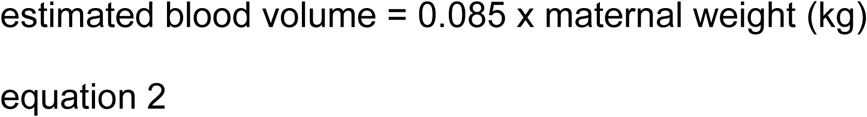

CBL is calculated blood loss in litres.

Hb_prebirth_ is haemoglobin measured shortly before birth

Hb_postpartum_ is postpartum haemoglobin typically measured 24 hours after birth.

For some women, 24-hour postpartum haemoglobin is higher than prebirth haemoglobin, resulting in a negative calculated blood loss. As blood loss cannot be negative, we replaced these negative values with zero.

Shook and colleagues’ calculated blood loss formula uses haematocrit rather than haemoglobin, reflecting the era when haematocrit was the more commonly reported measure in obstetric research.^15^ However, the two measures are closely correlated: haematocrit (as a percentage) is roughly 0.3 times the haemoglobin value in g/L, meaning that the proportional peripartum change is equivalent for either measure.^20,21^ Haemoglobin is now more commonly used in clinical practice, making it the more practical choice. We repeated key analyses by replacing haemoglobin with haematocrit to confirm that our results are robust to either measure (see Appendix 3).

### Statistical Analysis

We report median (IQR) measured and calculated blood loss using Wilcoxon signed-rank tests for comparison. We use Spearman’s correlation coefficient to quantify the association between measured and calculated blood loss. We use scatter graphs to show the relationship between measured and calculated blood loss for each woman, and Bland-Altman graphs to assess agreement and quantify bias. We report Cohen’s kappa to quantify agreement between measured and calculated blood loss at thresholds of 300, 500, 1000 and 1500 ml. We conduct these analyses for all women and then stratified by anaemia status. We used bootstrapping to test whether the Spearman correlation between measured and calculated blood loss was the same in women without anaemia and women with anaemia.

Because women with anaemia have a lower baseline haemoglobin concentration, a given volume of blood lost produces a smaller absolute drop in haemoglobin than in women without anaemia. However, calculated blood loss is based on the principle that the proportional drop should be the same in both groups. We use scatter graphs to show mean absolute and proportional haemoglobin drop by category of measured blood loss, stratified by anaemia status.

We report sensitivities, specificities, positive predictive values (PPV) and negative predictive values (NPV) to quantify the ability of different thresholds of measured and calculated blood loss to predict death or near-miss and haemodynamic compromise. We define death or near-miss as death from bleeding, transfusion with ≥ 5 units of blood, renal dysfunction (oliguria non-responsive to fluids or diuretics, dialysis for acute renal failure, or severe acute azotaemia), clotting failure, cardiovascular dysfunction (cardiac arrest, severe hypoperfusion, severe acidosis, or administration of continuous vasoactive drugs or cardiopulmonary resuscitation), hysterectomy, laparotomy to control bleeding, or arterial embolization or ligation. Haemodynamic compromise was defined as postpartum shock index >1.0, where shock index is heart rate divided by systolic blood pressure. Shock index was originally described by Allgöwer and colleagues, who reported that values around 1.0 indicate impending shock, and has been evaluated as a predictor of adverse outcomes in maternal haemorrhage.^22,23^ We show receiver operating characteristic (ROC) curves and report the area under the curve (AUC) to summarise the ability of measured and calculated blood loss to predict both outcomes. We use DeLong’s test to calculate p-values for differences in AUC.

## Results

### Study population

As of 11 May 2026, the I’M WOMAN trial had recruited 29,849 women. Of these, 10,468 women having a caesarean birth were recruited after 27 June 2025 from 32 hospitals where the protocol amendment requiring measurement of prebirth and postpartum haemoglobin and weight had been implemented. Table 1 shows the characteristics of our study population. Women were recruited from Nigeria (29%), Pakistan (36%) and Tanzania (35%). Their median (IQR) age was 30 (26-34) years, with 63% of women undergoing an emergency caesarean and 37% a planned caesarean. Median time from caesarean skin closure to postpartum haemoglobin measurement was 23 (23-24) hours. Weight was measured prebirth for 78% of women and estimated for the remaining 22%. We had the following missing values: prebirth haemoglobin (31), postpartum haemoglobin (45), weight (1). Missingness was low (<1%) and we conducted a complete case analysis.

**Table 1:** Patient characteristics of women included in this study. Participants are considered to have anaemia if their prebirth haemoglobin is less than 110 g/L. I’M WOMAN trial. N=10468.

| Variable | Number | Percent |
| --- | --- | --- |
| **Country** |  |  |
| Nigeria | 3018 | 28.8 % |
| Pakistan | 3816 | 36.5 % |
| Tanzania | 3634 | 34.7 % |
| **Mother's age (Years)** |  |  |
| Median (IQR) | 30 | (26-34) |
| <20 | 251 | 2.4 % |
| 20-29 | 4765 | 45.5 % |
| 30-39 | 4967 | 47.4 % |
| 40+ | 485 | 4.6 % |
| **Prebirth haemoglobin (g/L)** |  |  |
| Median (IQR) | 113 | (103-122) |
| No anaemia | 6182 | 59.1 % |
| Anaemia | 4255 | 40.6 % |
| Missing | 31 | 0.3 % |
| **Mother's Weight (Kg)** |  |  |
| Median (IQR) | 73 | (65-83) |
| <60 | 1111 | 10.6 % |
| 60-69 | 2787 | 26.6 % |
| 70-79 | 3073 | 29.4 % |
| 80+ | 3496 | 33.4 % |
| Missing | 1 | 0.0 % |
| **Weight status** |  |  |
| Measured | 8182 | 78.2 % |
| Estimated | 2286 | 21.8 % |
| **Gestational age (weeks)** |  |  |
| Median (IQR) | 38 | (37-39) |
| **Number of fetuses** |  |  |
| 1 | 10010 | 95.6 % |
| 2+ | 458 | 4.4 % |
| **Parity (includes this pregnancy)** |  |  |
| 1 | 2570 | 24.6 % |
| 2-4 | 6771 | 64.7 % |
| 5+ | 1127 | 10.8 % |
| **Units of blood transfused** |  |  |
| 0 | 9202 | 87.9 % |
| 1 | 900 | 8.6 % |
| 2 | 283 | 2.7 % |
| 3 | 45 | 0.4 % |
| 4 | 22 | 0.2 % |
| 5+ | 16 | 0.2 % |
| **Caesarean type** |  |  |
| Emergency | 6552 | 62.6 % |
| Planned | 3916 | 37.4 % |
| **Time from closure to FBC (hours)** |  |  |
| Median (IQR) | 23 | (23-24) |

Denominators vary between analyses because complete case analysis was used. We
had the following missing values: prebirth haemoglobin (31), postpartum haemoglobin
(45), weight (1).

### Comparing measured and calculated blood loss

Figure 1 and Table 2a summarise blood loss by each measurement method. Median (IQR) blood loss was 545 (430–700) ml for measured blood loss and 505 (180–908) ml for calculated blood loss (p<0.001). The IQR was wider for calculated blood loss than for measured blood loss, indicating greater variability in calculated blood loss. Calculated blood loss was negative for 1463 (14%) women before replacement with zero. Negative calculated blood loss values were replaced with zero for all analyses except Figure 1, which shows the unmodified distribution. The monotonic association between measured and calculated blood loss was weak (rho=0.28). A scatter plot (Appendix 2 Figure 1) shows considerable dispersion around the line of identity, indicating poor agreement between the two measures. Bland-Altman analysis (Figure 2) shows a small mean bias of -39 ml but wide limits of agreement (lower -1196 ml, upper 1118 ml). Systematic bias was present, with calculated blood loss exceeding measured blood loss as the mean of the two values increased. The number of women with blood loss above each threshold was: ≥ 300 ml: 9860 (95%) measured vs 6884 (66%) calculated; ≥ 500 ml: 6270 (60%) measured vs 5244 (51%) calculated; ≥ 1000 ml: 733 (7%) measured vs 2202 (21%) calculated; ≥ 1500 ml: 159 (2%) measured vs 867 (8%) calculated. Cohen’s kappa ranged from 0.03 at ≥ 300 ml to 0.24 at ≥ 1000 ml, indicating weak agreement. We repeated these analyses substituting haematocrit for haemoglobin when computing calculated blood loss and found that this made little difference to our results (see Appendix 3).

**Figure 1:**
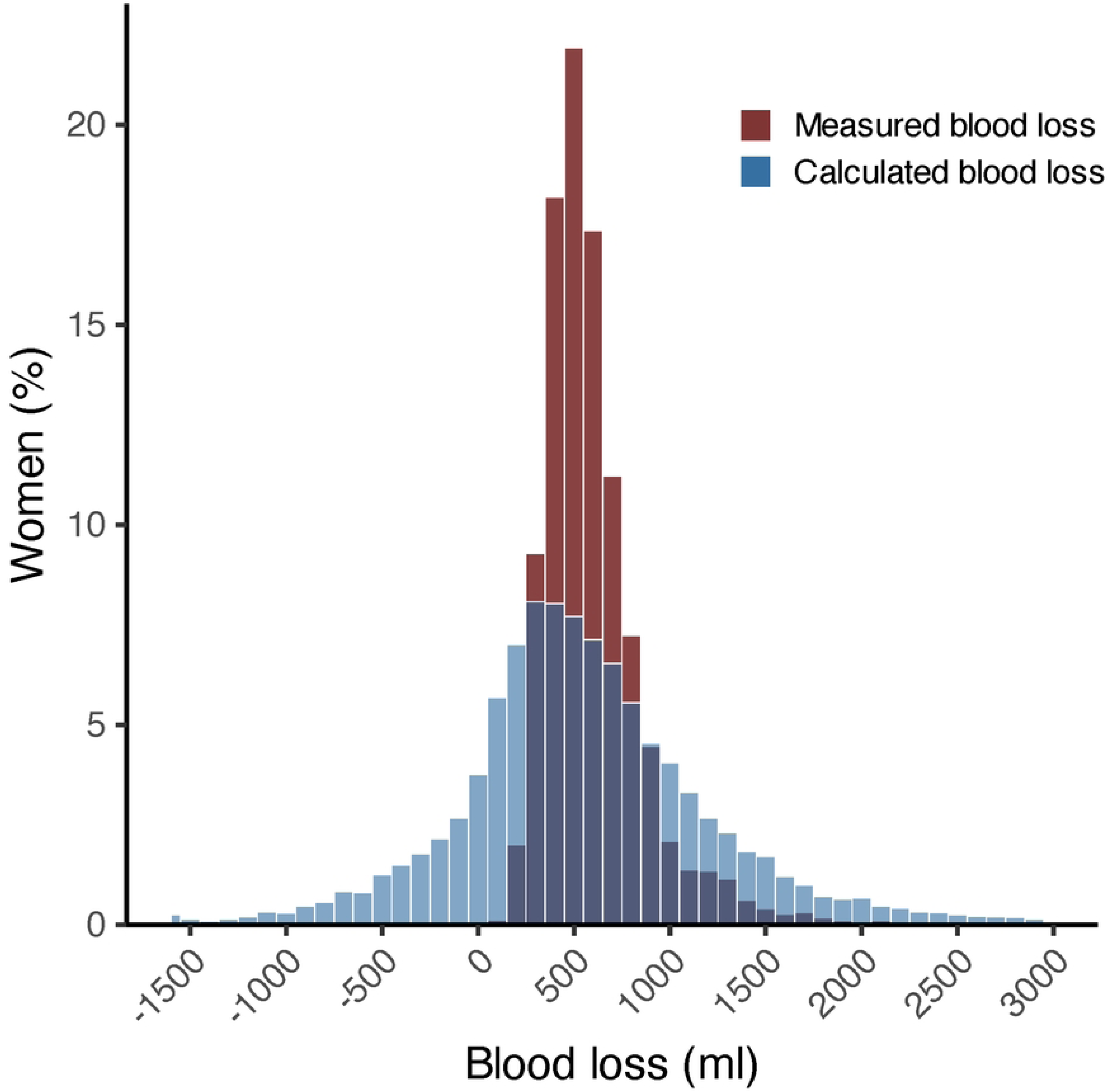
Histogram of Measured and Calculated blood loss. Median (IQR) blood loss is: Measured blood loss (545 (430–700) ml), Calculated blood loss (505 (180–908) ml). I’M WOMAN trial participants. N=10393. Negative calculated blood loss values are included in this graph. In all other analyses, tables figures and graphs, negative calculated blood loss values are replaced with zero.

**Figure 2:**
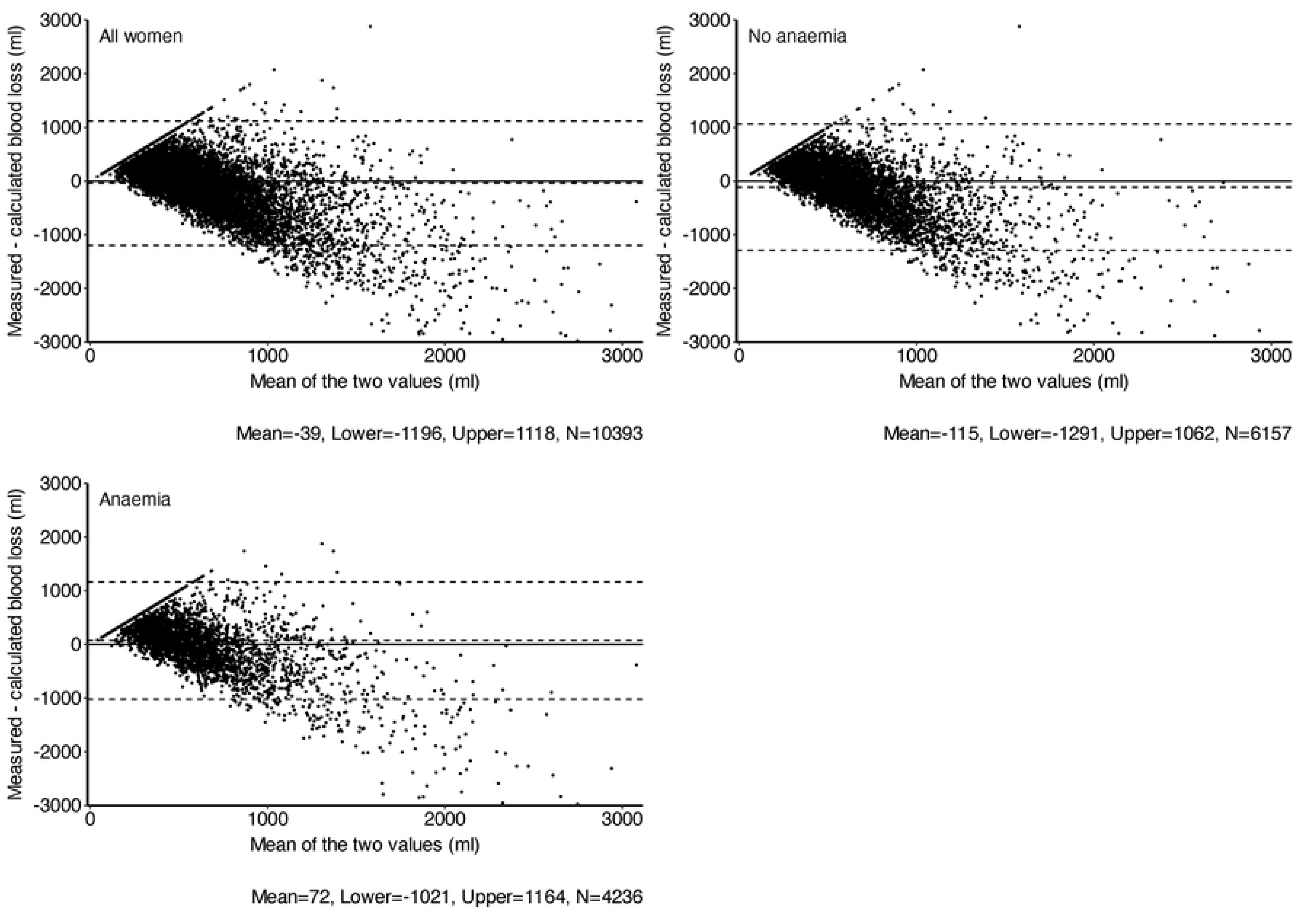
Bland-Altman plots of measured versus calculated blood loss. I’M WOMAN trial, Negative values of calculated blood loss have been replaced with zero.

**Table 2a:** Comparison of measured and calculated blood loss in all women. MBL: measured blood loss; CBL: calculated blood loss. Cohen’s Kappa measures agreement between methods at each blood loss threshold. Negative values of calculated blood loss were replaced with zero. P-values comparing means were calculated using a t-test and medians using a Wilcoxon signed-rank test.

| Measure | MBL | CBL | P-Value | Cohen's Kappa |
| --- | --- | --- | --- | --- |
| N | 10393 | 10393 |  |  |
| Mean (SD) | 605.5 (289.9) | 644.2 (659.2) | <0.001 |  |
| Median (IQR) | 545.0 (430.0–700.0) | 505.0 (180.0–908.0) | <0.001 |  |
| Range | 81 - 5600 | 0 - 7291 |  |  |
| Blood loss $\geq 300$, N (%) | 9860 (94.9%) | 6884 (66.2%) | | 0.034 |
| Blood loss $\geq 500$, N (%) | 6270 (60.3%) | 5244 (50.5%) | | 0.176 |
| Blood loss $\geq 1000$, N (%) | 733 (7.1%) | 2202 (21.2%) | | 0.244 |
| Blood loss $\geq 1500$, N (%) | 159 (1.5%) | 867 (8.3%) | | 0.182 |

Mean haemoglobin drop increased with measured blood loss and was higher in women without anaemia than in women with anaemia across all blood loss categories (Appendix 2 Figure 2). Figure 3 shows that women without anaemia had a higher proportional haemoglobin drop than women with anaemia for each category of measured blood loss below 1000 ml. When we stratified by anaemia severity using categories of no anaemia (haemoglobin ≥110 g/L), mild anaemia (100-109 g/L) and moderate or severe anaemia (<100 g/L), the results were similar (Appendix 4).

**Figure 3:**
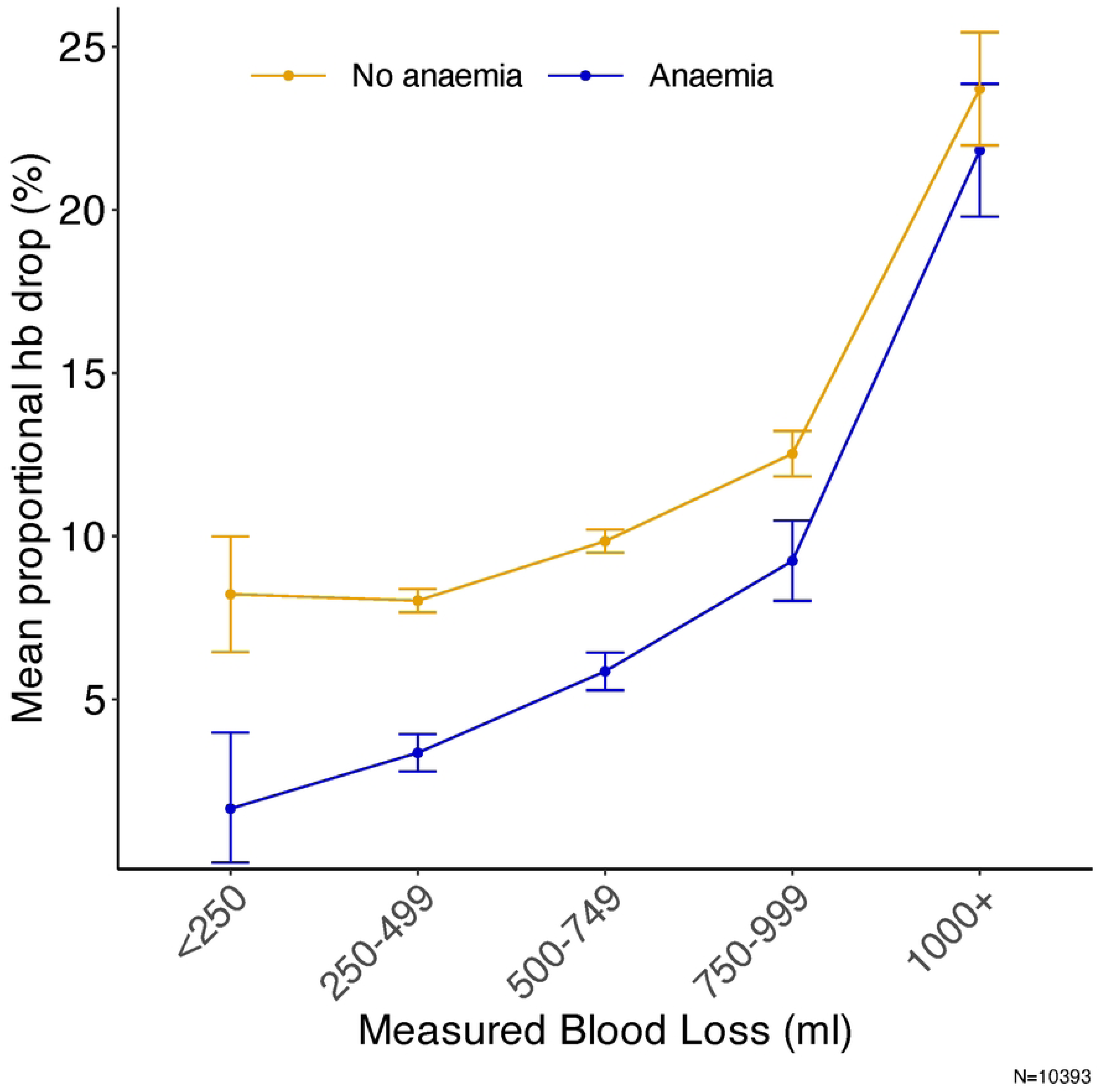
Measured blood loss vs mean proportional drop in haemoglobin stratified by anaemia status. I’M WOMAN trial. N=10,393. Anaemia was defined as prebirth haemoglobin < 110g/L. Negative values of calculated blood loss have been replaced with zero.

For women without anaemia, median (IQR) blood loss was 550 (431-700) ml for measured blood loss and 591 (272-997) ml for calculated blood loss (p<0.001; Table 2b). For women with anaemia, median (IQR) blood loss was higher for measured than calculated blood loss (541 (430-700) ml vs 378 (44-756) ml, p<0.001; Table 2c). The association between measured and calculated blood loss was weak in both women without anaemia and women with anaemia (rho=0.27 vs 0.30, p=0.25). For women without anaemia, Bland-Altman analysis (Figure 2) shows a negative mean bias of -115 ml (limits of agreement: -1291 ml to 1062 ml). For women with anaemia, the bias was 72 ml (limits of agreement: -1021 ml to 1164 ml). Table 2b shows that for women without anaemia, measured blood loss classified fewer women above higher thresholds than calculated blood loss (≥1000 ml: 7% vs 25%; ≥1500 ml: 2% vs 10%), with a similar pattern seen in women with anaemia (≥1000 ml: 8% vs 16%; ≥1500 ml: 1% vs 6%; Table 2c). Cohen’s kappa indicated poor to fair agreement between measured and calculated blood loss across all anaemia categories and blood loss thresholds (kappa 0.03-0.32).

**Table 2b:** Comparison of measured and calculated blood loss in women without anaemia. MBL: measured blood loss; CBL: calculated blood loss. Cohen’s kappa measures agreement between methods at each blood loss threshold. Negative values of calculated blood loss were replaced with zero. P-values comparing means were calculated using a t-test and medians using a Wilcoxon signed-rank test.

| Measure | MBL | CBL | P-Value | Cohen's Kappa |
| --- | --- | --- | --- | --- |
| N | 6157 | 6157 |  |  |
| Mean (SD) | 606.6 (291.9) | 721.2 (665.6) | <0.001 |  |
| Median (IQR) | 550.0 (431.0–700.0) | 591.0 (272.0–997.0) | <0.001 |  |
| Range | 135 - 5600 | 0 - 7291 |  |  |
| Blood loss $\geq 300$, N (%) | 5846 (94.9%) | 4498 (73.1%) | | 0.039 |
| Blood loss $\geq 500$, N (%) | 3748 (60.9%) | 3534 (57.4%) | | 0.169 |
| Blood loss $\geq 1000$, N (%) | 417 (6.8%) | 1534 (24.9%) | | 0.208 |
| Blood loss $\geq 1500$, N (%) | 104 (1.7%) | 595 (9.7%) | | 0.171 |

**Table 2c:** Comparison of measured and calculated blood loss in women with anaemia. MBL: measured blood loss; CBL: calculated blood loss. Cohen’s kappa measures agreement between methods at each blood loss threshold. Negative values of calculated blood loss were replaced with zero. P-values comparing means were calculated using a t-test and <u>medians using a Wilcoxon signed-rank test.</u>

| Measure | MBL | CBL | P-Value | Cohen's Kappa |
| --- | --- | --- | --- | --- |
| N | 4236 | 4236 |  |  |
| Mean (SD) | 603.9 (287.2) | 532.3 (633.4) | <0.001 |  |
| Median (IQR) | 541.0 (430.0–700.0) | 378.0 (43.5–756.0) | <0.001 |  |
| Range | 81 - 4150 | 0 - 7140 |  |  |
| Blood loss $\geq 300$, N (%) | 4014 (94.8%) | 2386 (56.3%) | | 0.029 |
| Blood loss $\geq 500$, N (%) | 2522 (59.5%) | 1710 (40.4%) | | 0.182 |
| Blood loss $\geq 1000$, N (%) | 316 (7.5%) | 668 (15.8%) | | 0.319 |
| Blood loss $\geq 1500$, N (%) | 55 (1.3%) | 272 (6.4%) | | 0.203 |

### Comparing the ability of measured and calculated blood loss to predict adverse outcomes

Table 3a and Figure 4a show the performance of measured and calculated blood loss in predicting death or near-miss. Death or near-miss occurred in 1.0% (106/10393) of women. Measured blood loss showed good performance in predicting death or near-miss (AUC=0.87). For a 300 ml threshold, sensitivity was 100% but specificity was only 5% and PPV only 1%. Higher thresholds improved specificity (94% at 1000 ml; 99% at 1500 ml) but reduced sensitivity (70% and 41% respectively). PPV was 27% for a 1500 ml threshold. NPV was 99% or higher across thresholds. The AUC for calculated blood loss (0.76) was lower (p<0.001) than for measured blood loss. For a 300 ml threshold, sensitivity was 84% and specificity was 34%. At 1500 ml, sensitivity was 56% but PPV was only 7%. NPV was 100% across all thresholds (300 ml, 500 ml, 1000 ml, 1500 ml).

**Table 3a:** Predictive value of measured and calculated blood loss for death or near-miss. Death or near-miss occurred in 106 women. We define death or near-miss as death from bleeding, transfusion with ≥5 units of blood, hysterectomy, laparotomy to control bleeding, arterial embolization or ligation, renal dysfunction, cardiovascular dysfunction or clotting failure. PPV is positive predictive value. NPV is negative predictive value. Negative values of calculated blood loss were replaced with zero. I’M WOMAN trial. N=10393.

| Predictor | Threshold (ml) | PPV (%) | NPV (%) | Sensitivity (%) | Specificity (%) | AUC |
| --- | --- | --- | --- | --- | --- | --- |
| Measured blood loss | 300 | 1 | 100 | 100 | 5 | 0.87 |
|  | 500 | 2 | 100 | 93 | 40 |  |
|  | 1000 | 10 | 100 | 70 | 94 |  |
|  | 1500 | 27 | 99 | 41 | 99 |  |
| Calculated blood loss | 300 | 1 | 100 | 84 | 34 | 0.76 |
|  | 500 | 2 | 100 | 76 | 50 |  |
|  | 1000 | 3 | 100 | 67 | 79 |  |
|  | 1500 | 7 | 100 | 56 | 92 |  |

**Figure 4a:**
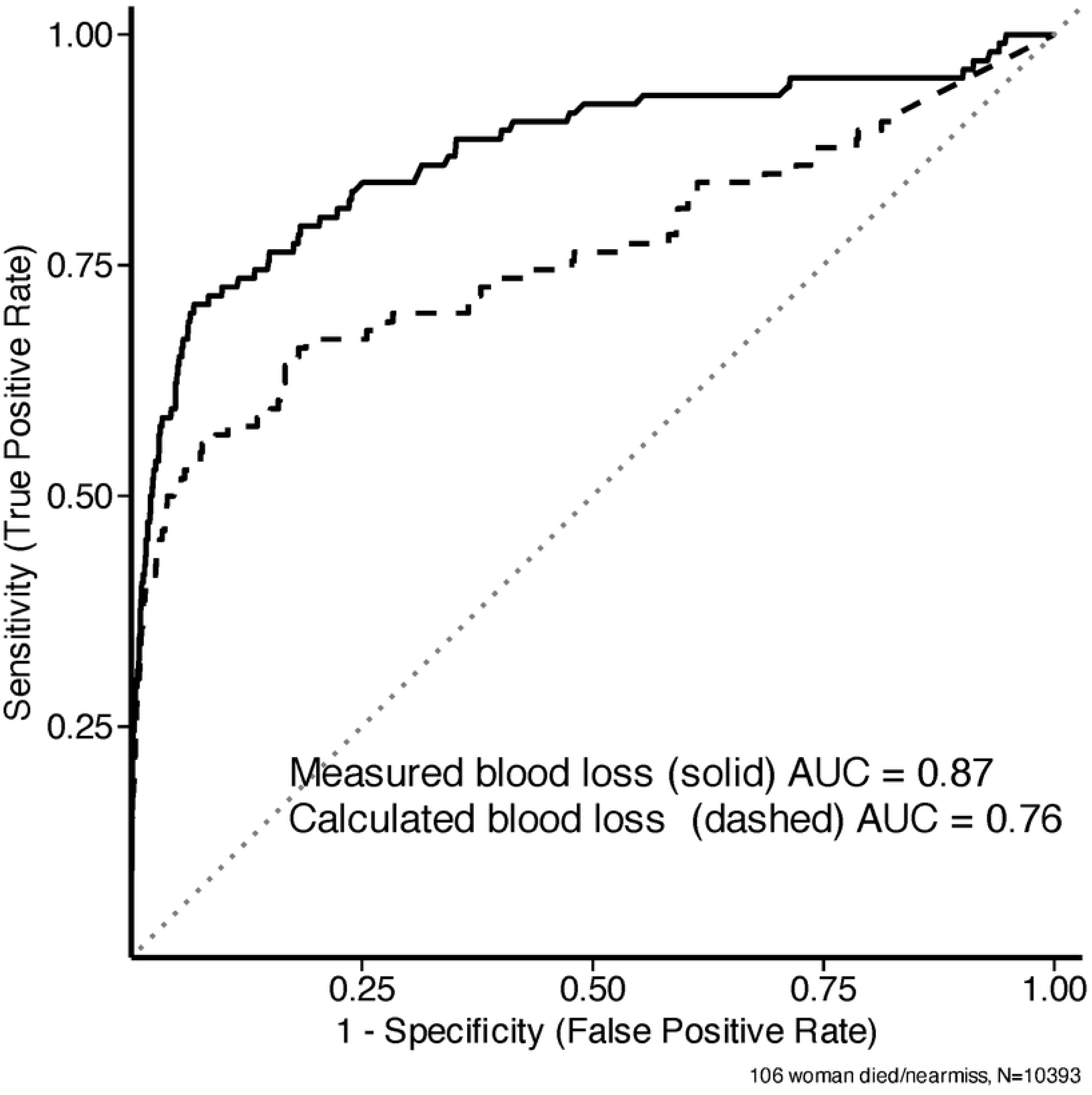
ROC curve for measured blood loss and calculated blood loss in predicting death or near miss. I’M WOMAN trial N=10393. Death or near-miss includes death from bleeding, transfusion with ≥5 units of blood, hysterectomy, laparotomy to control bleeding, arterial embolization or ligation, renal dysfunction, cardiovascular dysfunction or clotting failure. Negative values of calculated blood loss have been replaced with zero.

Table 3b and Figure 4b show the performance of measured and calculated blood loss in predicting haemodynamic compromise. Haemodynamic compromise occurred in 8.3% (862/10393) of women. Measured blood loss showed fair performance in predicting haemodynamic compromise (AUC=0.66). At a threshold of 300 ml, sensitivity was high (96%) but specificity was poor (5%) and PPV was only 8%. Higher thresholds improved specificity (95% at 1000 ml; 99% at 1500 ml) but reduced sensitivity (25% and 7% respectively). NPVs were above 90% across all thresholds (300 ml, 500 ml, 1000 ml, 1500 ml). The AUC for calculated blood loss was 0.62, lower than for measured blood loss (p=0.001). Sensitivity at 300 ml was 75%, lower than for measured blood loss at the same threshold, with a specificity of 35%. At 1000 ml, sensitivity was 39% and specificity was 80%; at 1500 ml, sensitivity was 20% and specificity was 93%. PPVs were low across all thresholds (9–20%), and NPVs were between 93% and 94% throughout. Measured blood loss was a better predictor of haemodynamic compromise than calculated blood loss across a range of shock index thresholds (SI >0.9 to >1.2; Appendix 5, table 1). There was strong evidence of a difference in predictive ability at each threshold (p≤0.003 in each case). At higher thresholds (SI >1.3 and >1.4), the difference in AUC remained similar in magnitude but the evidence was weaker, likely reflecting the small number of events (101 and 59, respectively). We report the predictive ability of measured and calculated blood loss for dizziness, nausea and vomiting in Appendix 5, tables 2-4. For all outcomes, measured blood loss had a modestly higher AUC than calculated blood loss.

**Table 3b:** Predictive value of measured and calculated blood loss for haemodynamic compromise. Haemodynamic compromise occurred in 862 women. We define haemodynamic compromise as postpartum shock index > 1.0, where shock index = heart rate ÷ systolic blood pressure. PPV is positive predictive value. NPV is negative predictive value. AUC is area under the curve. Negative values of calculated blood loss were replaced with zero. I’M WOMAN trial. N=10393.

| Predictor | Threshold (ml) | PPV (%) | NPV (%) | Sensitivity (%) | Specificity (%) | AUC |
| --- | --- | --- | --- | --- | --- | --- |
| Measured blood loss | 300 | 8 | 94 | 96 | 5 | 0.66 |
|  | 500 | 10 | 95 | 76 | 41 |  |
|  | 1000 | 29 | 93 | 25 | 95 |  |
|  | 1500 | 36 | 92 | 7 | 99 |  |
| Calculated blood loss | 300 | 9 | 94 | 75 | 35 | 0.62 |
|  | 500 | 11 | 94 | 65 | 51 |  |
|  | 1000 | 15 | 94 | 39 | 80 |  |
|  | 1500 | 20 | 93 | 20 | 93 |  |

**Figure 4b:**
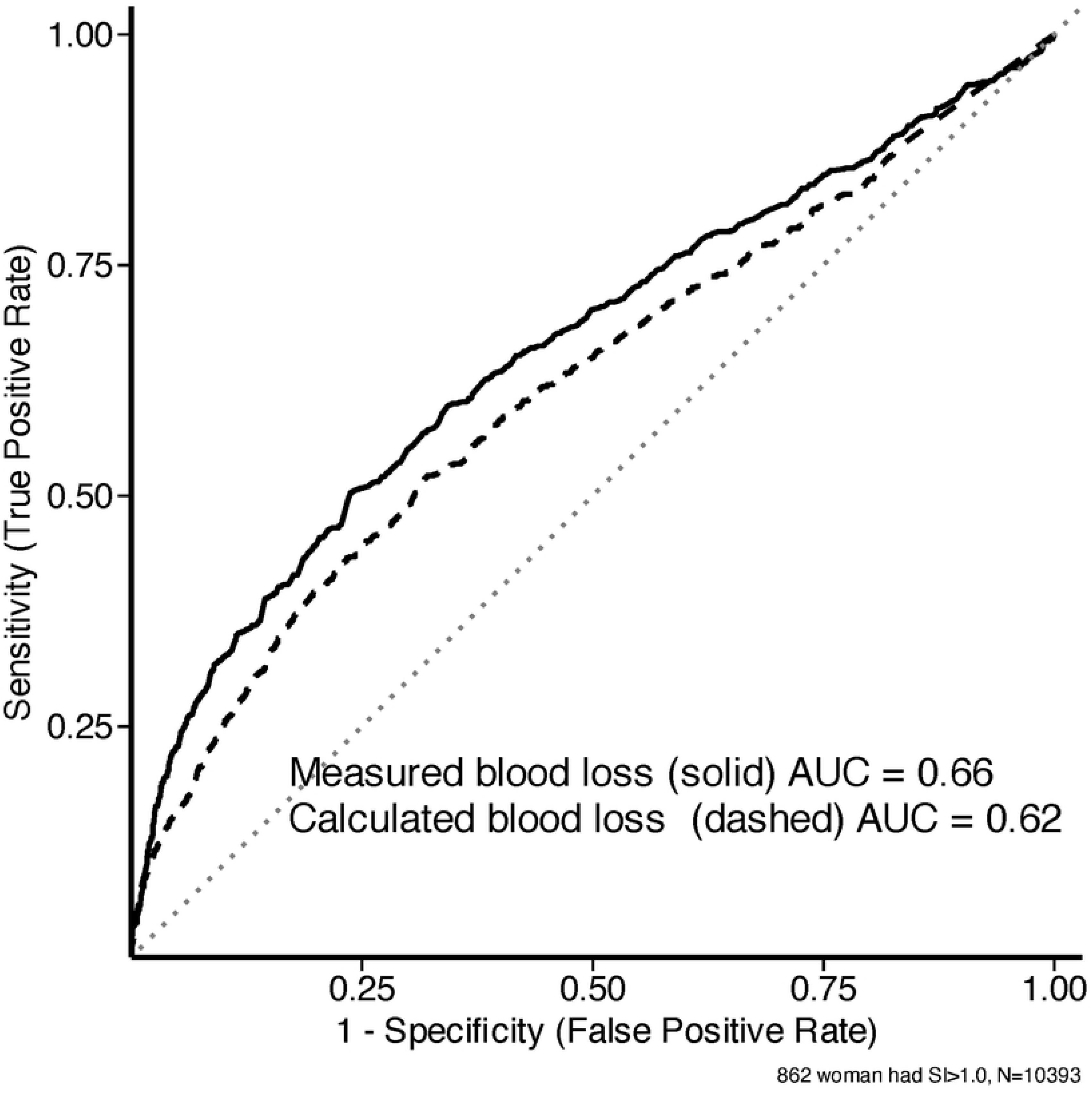
ROC curve for Measured blood loss and Calculated blood loss in predicting haemodynamic compromise. I’M WOMAN trial N=10393. We define haemodynamic compromise as postpartum shock index above 1.0 where shock index = heart rate ÷ systolic blood pressure. Negative values of calculated blood loss have been replaced with zero.

## Discussion

Median measured and calculated blood loss were similar overall. However, calculated blood loss classified more women as having clinically important blood loss: three times as many had blood loss above 1000 ml and five times as many had blood loss above 1500 ml. Cohen’s kappa indicated fair agreement at the 1000 ml threshold and poor agreement at the 1500 ml threshold. The association between measured and calculated blood loss was weakly monotonic. Measured blood loss was modestly better than calculated blood loss at predicting death or near-miss and haemodynamic compromise.

The association between measured blood loss and proportional peripartum haemoglobin drop was larger in women without anaemia than in women with anaemia for blood losses below 1000 ml. Since calculated blood loss is a linear function of proportional peripartum haemoglobin drop, this may explain why median calculated blood loss was higher than measured blood loss in women without anaemia but lower in women with anaemia.

Both measured and calculated blood loss are imperfect measures. Clinicians caring for women in this study reported that measured blood loss is often underestimated due to blood spilled during surgery or lost outside the drape. In addition, when a woman is bleeding heavily, clinicians might stop measuring and prioritise treatment. Measured blood loss may also be overestimated due to contamination with amniotic and surgical irrigation fluid. Calculated blood loss assumes that a woman’s circulating blood volume is the same at the time of her prebirth and postpartum haemoglobin measurements. However, blood volume increases during pregnancy and falls in the first 24 hours after birth, violating this assumption.^24,25^ Some women receive intravenous fluids during and after birth, diluting their blood. This dilution reduces the postpartum haemoglobin measurement and increases calculated blood loss regardless of the actual amount of blood lost.

Despite its modestly lower predictive ability, calculated blood loss has advantages. It only requires three routinely collected measures (maternal weight and prebirth and postpartum haemoglobin), making it practical in settings where the equipment and trained staff needed for measured blood loss are unavailable. In addition, it is not subject to mismeasurement due to spillage or contamination with amniotic and irrigation fluid, and its standardised nature makes it a suitable outcome measure for clinical trials.

Tan and colleagues compared measured and calculated blood loss in 197 women undergoing a planned caesarean section in the largest maternity unit in Singapore.^4^ Measured blood loss was assessed by combining the weighing of blood-soaked surgical swabs and postpartum drapes with the volume of blood in the suction canister. Blood volume in the canister was isolated by subtracting the recorded volumes of amniotic fluid and irrigants from the total fluid collected. Their methods for obtaining calculated blood loss were similar to ours. Preoperative haematocrit was measured from venous blood within one week before surgery, with a repeat measurement taken 48 hours after birth. They found that median measured blood loss was lower than calculated blood loss, although there was no statistical evidence of a difference, and that the two measures were only weakly correlated.

Madar and colleagues compared measured and calculated blood loss in 8341 women having an in-hospital vaginal birth in France.^9^ Measured blood loss was obtained from calibrated drapes. Calculated blood loss was obtained using methods similar to ours. Haematocrit was measured in the eighth or ninth month of pregnancy and 48 hours after birth. They found that median measured blood loss was lower than median calculated blood loss, with moderate correlation between the two measures.

Key strengths of this study include our large sample size and consistent haemoglobin measurement protocol. Haemoglobin was measured from venous samples and all women had their haemoglobin measured after hospital arrival but before birth. Postpartum haemoglobin was measured more than 12 hours after birth in over 99% of women, and more than 22 hours after birth in 95%. This timing is important as previous studies show that postpartum haemoglobin falls significantly after 12 hours and approaches its lowest level by 24 hours.^26^ When we repeated key analyses replacing haemoglobin with haematocrit, our results were similar, suggesting that our findings are robust to the choice of measure.

A limitation of our study is the absence of a gold standard measure of blood loss. Ideally, measured blood loss would have been validated against colorimetric or spectrophotometric laboratory methods in a subset of our study population.^27–29^ Validation would have allowed us to correct for contamination with amniotic and surgical irrigation fluid. We did not have data on intravenous fluids given to women in this study, so could not correct for haemodilution, though in practice few studies correct for intravenous fluid administration when calculating blood loss. In a previous obstetric trial of 15,066 women having an in-hospital vaginal birth in Nigeria, Pakistan, Tanzania and Zambia, we found that 30% of women received fluid intravenously.^19^ Site monitoring suggests that most of the women in this study received intravenous fluids. A further limitation is that despite our large overall sample size, the number of women who experienced death or near-miss was small.

## Conclusions

Calculated blood loss classified three times as many women as having blood loss above 1000 ml compared with measured blood loss, and the two measures were only weakly correlated. Measured blood loss had a modest advantage in predicting death or near-miss and haemodynamic compromise, but the objective nature of calculated blood loss makes it a suitable outcome for clinical trials when direct measurement is not feasible.

## Data Availability

After publication of primary and secondary analyses detailed in the statistical analysis plan, individual de-identified patient data, including the data dictionary, will be made available via our data sharing portal, The Free Bank of Injury and Emergency Research Data (FreeBIRD), indefinitely to allow maximum use of the data to improve patient care and advance medical knowledge. https://freebird.lshtm.ac.uk

## Author contributions

RM and IR planned the analyses. RM drafted the manuscript with help from IR, MA and KK and with input from all authors. RM did the statistical analyses, interpreted the data, and generated the figures. RC was lead coordinating investigator in Pakistan. NB was lead coordinating investigator in Nigeria. PM was lead coordinating investigator in Tanzania. MA, KK, DP and AG interpreted the data. AG and DP managed the data collection. RM, DP, KK, MA and AG had direct access to the data and verified the data reported in the manuscript. All authors were involved in the development of the original manuscript, and have read and approved the final version submitted.

